# Predicted Symptomatic Effectiveness of Pfizer-BioNTech BNT162b2 Vaccine Against Omicron Variant of SARS-CoV-2

**DOI:** 10.1101/2021.12.09.21267556

**Authors:** Oleg Volkov

## Abstract

This paper presents predictions of the symptomatic effectiveness of the Pfizer-BioNTech BNT162b2 (Comirnaty) vaccine against Omicron B.1.1.529, the latest SARS-CoV-2 variant of concern. They were obtained assuming fold decreases in Omicron neutralisation by vaccine-induced antibodies versus neutralisation of the virus Wild Type. A 25-fold decrease was assumed based on Omicron pseudovirus neutralisation study by Pfizer and BioNTech; a 94-fold, based on live-Omicron neutralisation study in South Africa; and 40, 80 and 120 folds, hypothesised based on genetic information. The effectiveness of two vaccine doses was predicted as 66% (42, 86), 48% (25, 72) and 42% (20, 66) for up to five months starting 2-4 weeks after the second dose, for the 25, 80 and 120 folds, respectively. The effectiveness of booster vaccination was predicted under a highly conservative assumption that the third dose would increase neutralisation by only 3.3 folds compared to the second dose. The predictions of effectiveness for up to five months, starting 2-4 weeks after the third dose, were 81% (59, 95), 67% (43, 87) and 61% (37, 82) for the 25, 80 and 120 folds, respectively. Despite the large fold decreases considered, the vaccine could still provide substantial protection, particularly after a booster and against severe disease. The paper is accompanied by free software which can be used to predict the symptomatic effectiveness of Comirnaty against Omicron under different neutralisation folds, including those obtained experimentally.

---

The latest SARS-CoV-2 B.1.1.529 Omicron variant of concern (VOC) has uncharacteristically many mutations heightening the risk of immune escape. There is an exceptional urgency to assess how well vaccine protection would hold up. We predicted the symptomatic effectiveness of the Pfizer-BioNTech BNT162b2 (Comirnaty) vaccine based on experimental results from the vaccine developers [Pfizer/BioNTech, 2021] and from researchers in South Africa [Cele et al., 2021]. These experiments gave early estimates of Omicron neutralisation by vaccine-induced antibodies, compared to neutralisation of the virus Wild Type. Since these results are preliminary and more studies are forthcoming, we also made predictions under hypothetical scenarios for reduced neutralisation of this VOC.

The predictive framework used and its accompanying software can be valuable for assessing other neutralisation scenarios and to relating neutralisation study results to Comirnaty effectiveness against Omicron. The framework was developed in Volkov et al. [2021] by building upon Khoury et al. [2021]. It models the effectiveness of Covid-19 vaccines as a function of 50% neutralising antibody titres against SARS-CoV-2 variants. Its predictions for Omicron used an effectiveness model fitted to 32 estimates from symptomatic effectiveness studies with the vaccine and to neutralising antibody data on the vaccine from the Crick/UCLH Legacy study [Wall et al., 2021] with previous VOCs. The framework assumptions, caveats and methodology are summarised in the appendix and in Volkov et al. [2021].

## 1 Results

The neutralisation scenarios behind the predictions are defined in Section A.1. Each scenario specifies a fold decrease of Omicron neutralisation compared to neutralisation of the SARS-CoV-2 Wild Type. The fold decreases are also stated versus neutralisation of Delta after two Comirnaty doses from the Legacy Study [Wall et al., 2021]. The latter folds were used to construct the neutralising titres assumed for Omicron; see Section A.2.

### Full primary vaccination (two doses)

We consider adult populations and assume that Omicron and previous variants cause similar symptoms. Effectiveness predictions after two Comirnaty doses were obtained for a period of 3-5 months starting 2-3 weeks after the second dose. (These assumptions reflect the effectiveness data used to fit the prediction model.) The Omicron versus Delta fold-decrease is defined as Omicron versus the Wild Type decrease divided by 5.8, the corresponding fold-decrease for Delta versus the Wild Type in the Legacy Study, for example, as 80*/*5.8 = 13.8 under the Polymutant scenario. The predictions, using the model in Section A.3 are listed in Table 1 and illustrated in Figure 1. The predicted effectiveness of 49.9% (27, 73), just below the FDA-recommended threshold of 50%, corresponded to a 72-fold decrease.

**Table 1:**
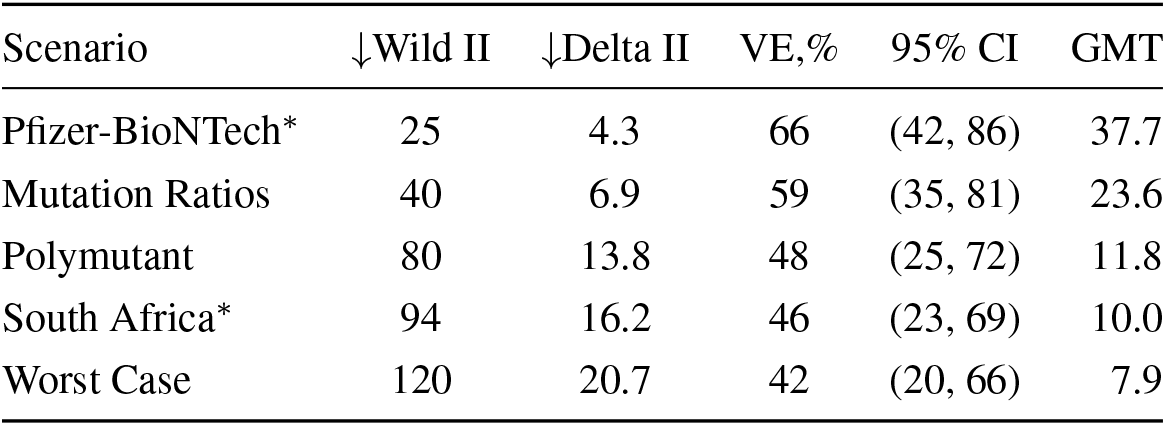
Predicted symptomatic effectiveness of full primary vaccination against Omicron. Fold decreases *↓* are defined for two doses of Comirnaty, versus Wild Type or Delta. The geometric mean titre (GMT) for the computed neutralising titres against Omicron is shown; see Section A.2. The hypothetical and the ^*\**^early-result scenarios are described in Section A.1.

**Figure 1:**
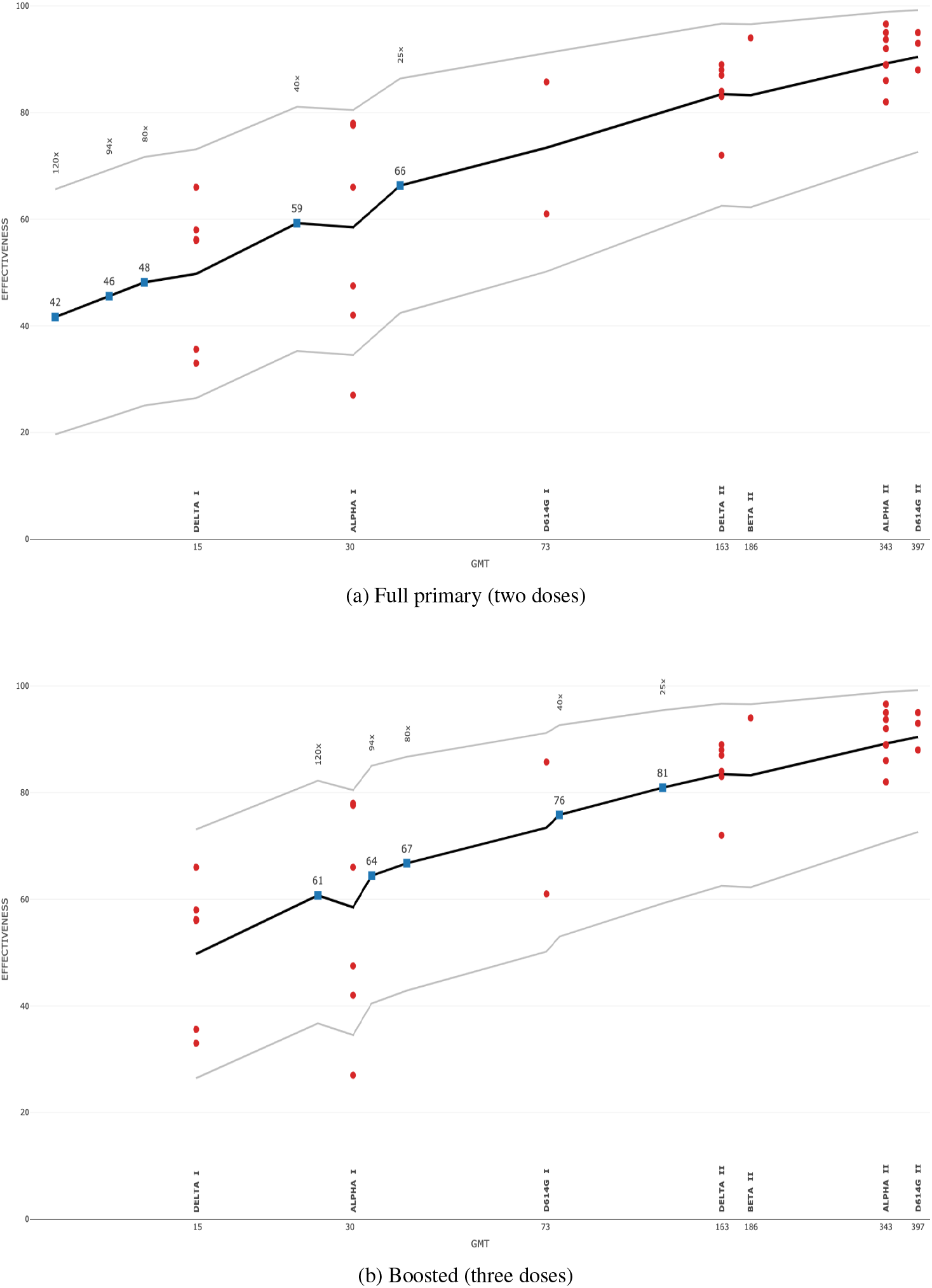
Predicted symptomatic effectiveness of Comirnaty vaccination against Omicron. The fitted model function (1) is plotted with 95% predictive bands. The vaccine doses and pre-Omicron variants are shown vs geometric mean titres for the Legacy Study and the estimates from effectiveness studies. The assumed fold-decreases from Omicron are shown vs predictions.

### Booster vaccination (three doses)

Besides the previous assumptions of symptom similarity and adult populations, we assume that a third Comirnaty dose is about 6 months after the second and consider 3-5 months starting 2-3 weeks after the third dose. Based on the approval documentation for Comirnaty boosters (Table 4 of FDA [2021]) we assumed that neutralisation a month after the third dose is 3.3 times that a month after the second. Other than that, the immune dynamics were assumed similar to those after the second dose. Since no neutralisation data were available on Comirnaty boosters, we computed Omicron titres after a booster as a fold of Delta titres, from the Legacy Study, after two vaccine doses. The corresponding fold was computed, for example, as 80*/*5.8*/*3.3 = 4.2 under the Polymutant scenario. The predictions are listed in Table 2 and illustrated in Figure 1. The effectiveness of 49.9% (27, 73), just below the FDA-recommended threshold of 50%, was predicted for 72*×*3.3 = 238 fold decreases with Omicron versus the Wild Type.

**Table 2:** Predicted effectiveness of booster vaccination against Omicron. Fold drop *↓* is defined versus 3 doses for Wild Type and 2 doses for Delta. The geometric mean titre (GMT) for the assumed distribution of neutralising titres is shown. The hypothetical and the ^*\**^early-result scenarios are described in Section A.1.

| Scenario | ↓Wild III | ↓Delta II | VE,% | 95% CI | GMT |
| --- | --- | --- | --- | --- | --- |
| Pfizer-BioNTech* | 25 | 1.3 | 81 | (59, 95) | 124.5 |
| Mutation Ratios | 40 | 2.1 | 76 | (53, 93) | 77.8 |
| Polymutant | 80 | 4.2 | 67 | (43, 87) | 38.9 |
| South Africa* | 94 | 4.9 | 64 | (40, 85) | 33.1 |
| Worst Case | 120 | 6.3 | 61 | (37, 82) | 25.9 |

### Geometric mean titres (GMT)

As described in Section A.2, the assumed neutralising titres against Omicron were computed as folds of the two-dose neutralising titres against Delta from the Legacy Study. For primary vaccination, every GMT of the corresponding Omicron titres was below 40, the lower detection limit in the Legacy study; see Table 1 and Figure 1. Starting with the Polymutant scenario, each GMT was below 15, the GMT against Delta in the one-dose cohort of the Legacy Study. Predictions far outside observed data, particularly under the Worst-Case scenario, may become increasingly less accurate. For boosted vaccination, each GMT was below 40 starting from the Polymutant scenario, but still above 15; see Table 2 and Figure 1. Such low titres against Omicron are illustrated in Figure 4 for primary vaccination under the Polymutant scenario: at least 90% of them are below the detection limit of 40. Such titres could be censored in practical studies, and hence provide little information. Neutralisation assays with Omicron may therefore require lower titres than the assay limits used with previous variants.

## 2 Discussion

The presented predictions are subject to uncertainties, for instance, due to population differences; see Section A.4. The fitted model did not incorporate the unavailable data on boosted vaccination for any VOC. Crucially, insufficient data currently exist on Omicron, both on its neutralisation folds relative to the Wild Type, and on the neutralising antibody titres against it. Once available, such data can be included into the model to enable more accurate predictions. Since the model used the rich data set from Legacy, additional data on Omicron and Comirnaty boosters from this study would greatly enhance the predictions. Data from other neutralisation studies, particularly whose assays include low titres, as well as from Comirnaty effectiveness studies, particularly in boosted vaccinees, would be also beneficial.

With the above caveats, we could make the following conclusions. Despite a 40-fold decrease in virus neutralisation, full primary vaccination with Comirnaty could be highly protective against Omicron, with the symptomatic effectiveness almost 60% for up to 5 months. Even with the Worst-Case scenario (120x decrease), two doses could still have a sizeable effectiveness of 42% (although note a potentially high prediction uncertainty at low titres). This implies that boosters might not need to be radically sped up for every population group. Protection after the third dose would be substantially stronger across all the scenarios considered, given the likely neutralisation increase relative to the second dose. In fact, the assumed 3.3-fold increase could be too conservative, given the 20x (15, 27) increase reported in Atmar et al. [2021] for homological Comirnaty boosting, and the 25x increase from the early data in Pfizer/BioNTech [2021] on Omicron pseudovirus neutralisation. The symptomatic effectiveness, predicted here for general adult populations, is likely to be higher in adolescents and young adults. The effectiveness against severe disease, hospitalisation and death is likely to exceed the predicted symptomatic effectiveness. If the early reports, such as [Chutel et al., 2021], that Omicron causes milder disease are confirmed, then the effectiveness could increase across the board.

Still, waning protection, especially after primary vaccination, will put a downward pressure on the effectiveness, particularly in vulnerable populations. The effectiveness of other vaccines, generating weaker immune responses, may drop to low levels. High transmissibility of Omicron may increase both vaccinated and unvaccinated cases, even if the vaccine effectiveness numbers hold up. These factors may necessitate early boosting in vulnerable populations, possibly with reduced doses to save vaccine quantity and prevent side effects. Immunity-boosting and dose-sparing heterological vaccination could also be warrantied, perhaps starting already from the second dose. The predictions also suggest that the Omicron impact on vaccine effectiveness is akin to giving at least one full vaccine dose less during the Alpha or Delta predominance. Even putting the ethics aside, this implies that vaccine stockpiling in some countries, while SARS-CoV-2 mutates unabated in others, is ultimately self-defeating.

## A Methods

### A.1 Neutralisation folds

To make predictions about Omicron, we would ideally use neutralising titres, against this VOC, from the Legacy or a compatible study. Since no such data are available, we assume fold-decreases for Omicron versus Wild Type titres. Predictions under any such folds — defined hypothetically or estimated experimentally — can be made with the provided software. Predictions in this paper are based on the following scenarios:

**Pfizer-BioNTech**\* 25x decrease

**Mutation Ratios** 40x decrease

**Polymutant** 80x decrease

**South Africa**\* 94x decrease

**Worst Case** 120x decrease

Either scenario^*^ is based on an early experimental result, such is that from a study by the Comirnaty developers [Pfizer/BioNTech, 2021]. This study tested neutralisation of the Omicron pseudovirus with sera taken from people three weeks after the second dose and from people one month after the third dose. It estimated a 25x fold decrease versus the Wild Type for the former, and also found that post-booster neutralisation against Omicron was comparable to two-dose neutralisation against “the ancestral strain” [Pfizer/BioNTech, 2021]. Therefore, booster effectiveness could be similar to the known effectiveness of two doses against the Wild Type, and hence no predictions would be required. The “Pfizer-BioNTech” scenario was defined with a 25x decrease, which corresponds to the study estimate for two doses but could be too pessimistic for three doses. (We assumed a 3.3x neutralisation increase after the third dose, rather than 25x suggested by the study.) The other early result came from a live-virus study of Omicron neutralisation in South Africa; Cele et al. [2021]. The fold-decrease was estimated in 12 people vaccinated with two Comirnaty doses, half of whom had had Covid-19 before. The estimate of 41.4x was given versus neutralisation of the D614G lineage, which in the Legacy Study had 2.3x (1.9, 2.6) lower neutralisation than the Wild Type. As a benchmark, we defined the “South Africa” scenario with 41.4 *×* 2.3 *≈* 94x decrease versus the Wild Type.

As the two estimates are quite different, and there is high uncertainty about Omicron neutralisation, we also examined three hypothetical scenarios, defined as follows. Their reference point was neutralisation folds for other VOCs, versus the Wild Type, assuming two Comirnaty doses; see Table 3. These folds came from live-virus neutralisation studies within the Legacy Wall et al. [2021], whose data were used to fit the prediction model, and also from ACI [2021], which based fold ranges on the WHO guidance about Comirnaty. Both sets of folds are consistent with the meta-analyses reported in Cromer et al. [2021] of Comirnaty neutralisation from six laboratories, including the Crick Institute in the Legacy Study. To interpret these folds, consider the progression of VOCs in the following three steps

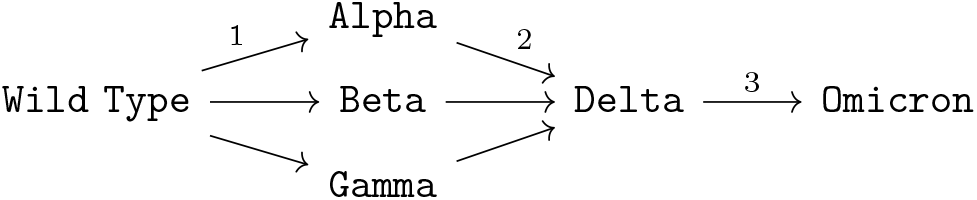

**Table 3:**
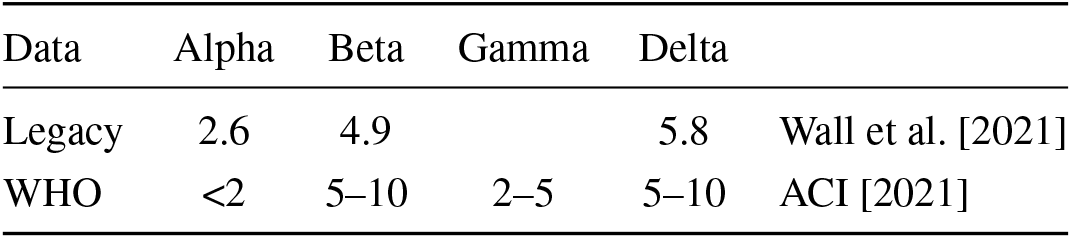
Neutralisation fold decreases vs Wild Type for two Comirnaty doses. The 95% CI for Legacy folds are (2.2, 3.1), (4.2, 5.7) and (5.0, 6.9), from left to right.

| Data | Alpha | Beta | Gamma | Delta |  |
| --- | --- | --- | --- | --- | --- |
| Legacy | 2.6 | 4.9 |  | 5.8 | Wall et al. [2021] |
| WHO | <2 | 5–10 | 2–5 | 5–10 | ACI [2021] |

The three initial VOCs emerged at the first step, with each overtaking an ancestor SARS-CoV-2 in a large geographical area: for instance, Beta in South Africa, Gamma in Brazil, and Alpha in most other countries; see covariants.org. At the second step, each of them was overtaken by Delta. The path to Delta via Beta suggests that a successor VOC need not be more resistant to neutralising antibodies (see Table 3), which is unrealistic for Omicron. The paths to Delta through Alpha and Gamma exhibited higher resistance to neutralisation after each step.

To define fold scenarios, we draw on what is already known about Omicron genetics. Table 4 is based on Bernasconi et al. [2021], with additional data from the Wikipedia pages on the VOCs. The displayed counts of Spike protein mutations and shares of affected B-cell epitopes are likely to be relevant to variant escape from neutralising antibodies. (The shares of affected T-cell epitopes were similar to those for B-cells in the table; Bernasconi et al., 2021.) The fold decreases, in Table 3, for the pre-Omicron variants appear correlated with the critical mutation counts. The relationship with the other two quantities is less clear — if anything, they are roughly the same for Delta, despite it overtaking the previous VOCs. All three quantities could still be indicative of Omicron neutralisation, given their dramatic hike for this VOC. Hence we might assess Omicron folds by combining the *𝒪/*Δ ratios from Table 4 with Delta folds from Table 3. The best-case prediction is 5 *×* 2.8 = 14x decreases versus the Wild Type; the worst case is 10 *×* 4.1 = 41x, which is set within the “Mutation Ratios” scenario. (The middle-range 25x fold is already covered by another scenario.)

**Table 4:**
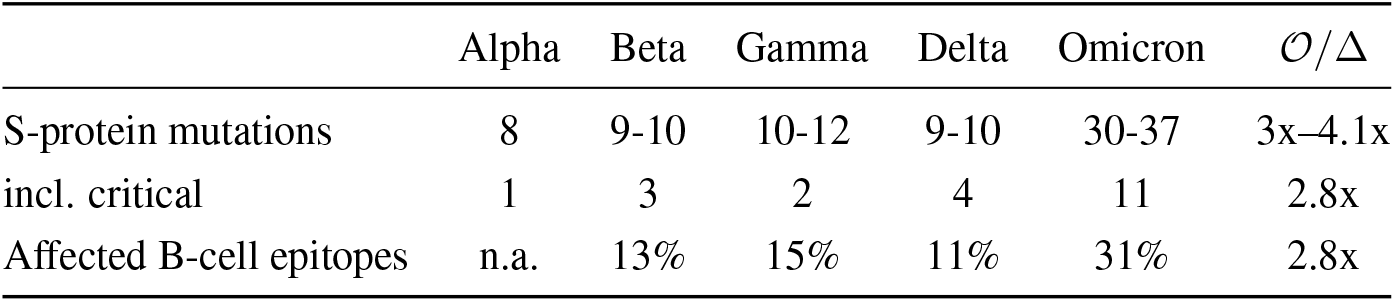
Omicron vs other VOCs. Main source: Bernasconi et al. [2021]; some mutation counts from Wikipedia. The last column shows rounded ratios for Omicron and Delta.

Given the unusually many mutations in Omicron, previous VOCs may be less informative than SARS-CoV-2 chimeric mutants synthesised to escape neutralising antibodies. A relevant benchmark could be the polymutant pseudovirus with 20 mutations deemed to maximise such escape [Schmidt et al., 2021], with many of its and Omicron’s mutations appearing in overlapping regions [Zimmer, 2021]. Schmidt et al. [2021] reported a 81-fold (range 8.4-229) decrease in polymutant neutralisation by sera of 14 Comirnaty-vaccinated people versus the Wild Type. There are reasons why the Omicron fold-decrease could be smaller. For instance, as noted in Schmidt et al. [2021], the polymuntant incurred fitness costs: a benchmark pseudovirus encoding the ancestral spike proteins replicated about 15 times faster; see Figure 3b of the paper. Conversely, there are reasons why the Omicron fold-decrease could exceed 81. For instance, this VOC had at least 37 mutations just on the Spike protein, and 20 or more elsewhere, compared to 20 overall for the polymutant. Another complication for gauging the fold-decrease is the uncertainty in its estimate, as there were only 14 subjects, and there could also have been systematic error. For instance, Figure 2 plots the estimated 50% neutralising titres, versus the Wild Type, for replicate 1 and replicate 2 in the study, based on the data for Figure 3 of Schmidt et al. [2021]. That all the titres for replicate 1 were 1.3–2.6 times higher than for replicate 2 suggests a possible systematic error. (No such trend appeared in the data on other variants and for convalescent subjects.) Error could also be due to the use of a pseudovirus, as what the polymutant essentially was. Even pseudoviruses created to maximally mimic natural SARS-CoV-2 viruses can have markedly different neutralisation to that of live virus by the same sera; see Figures 3 and 4 of Lu et al. [2021]; Wang et al. [2021]. To accommodate these uncertainties, we assumed that the actual Omicron fold could be within 80 *±* 40. Since 40x is already covered, we defined the “Polymutant” scenario with 80x and the “Worst-Case” scenario with 120x.

**Figure 2:**
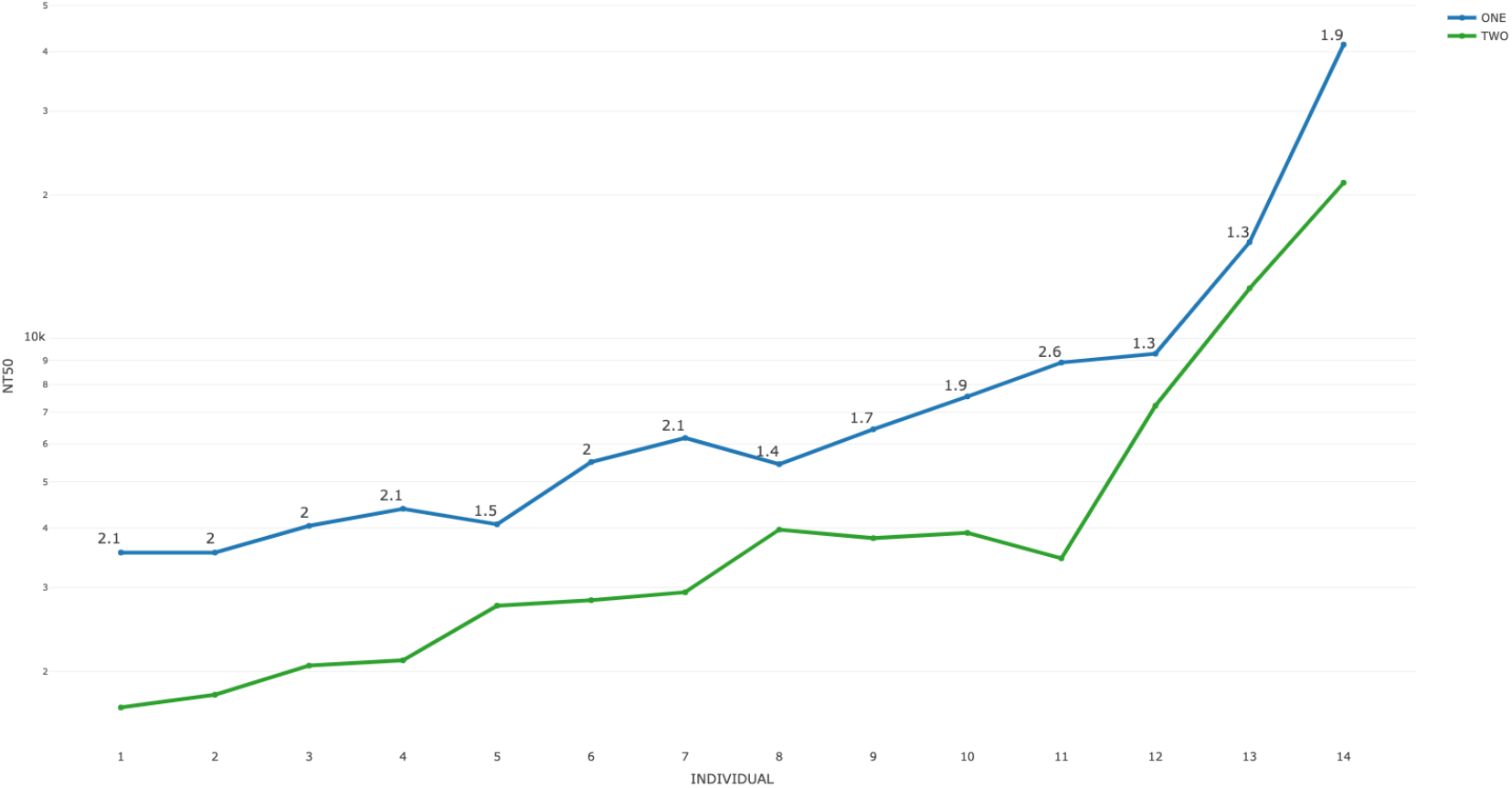
Log-transformed 50% neutralising titres vs Wild Type for 14 vaccinees for Replicate One and Two. The ratios One/Two are shown on the original scale. Source: Schmidt et al. [2021], data for Figure 3.

**Figure 3:**
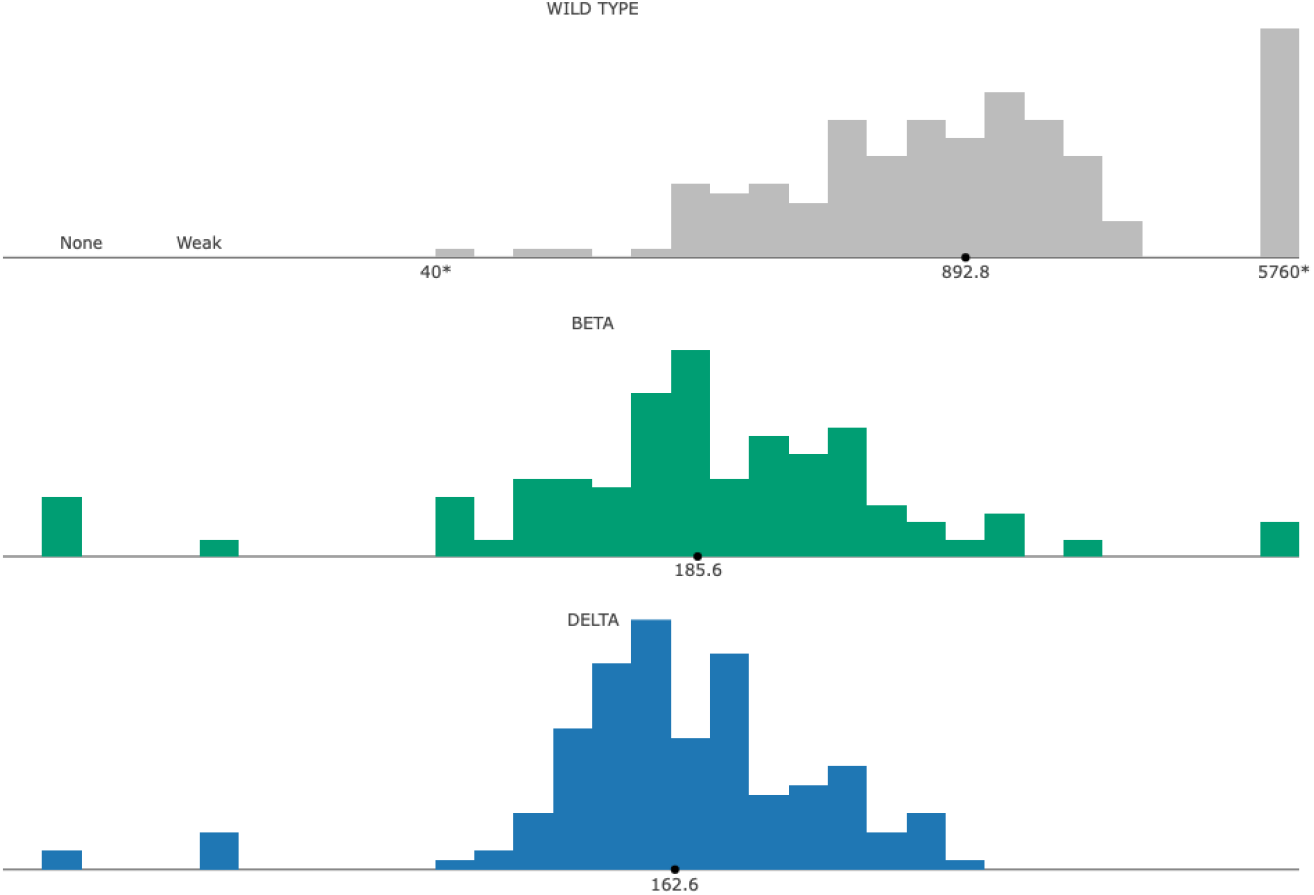
Log10-transformed 50% neutralising titres from the two-dose cohort of the Legacy Study. The GMTs are labeled on the natural scale.

**Figure 4:**
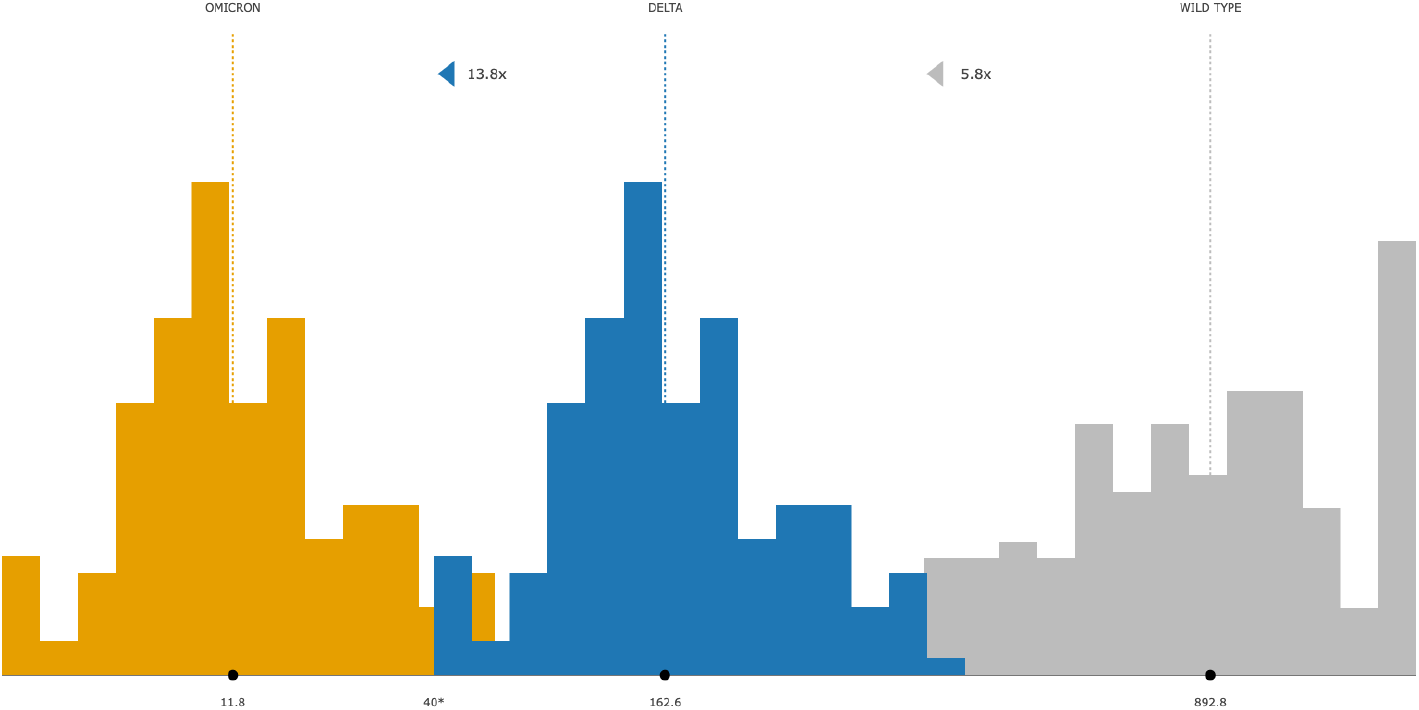
Log10 titres against Omicron after two doses for the Polymutant scenario with 80x. First is the 5.8x shift from the Wild Type to Delta. Then, the Delta titres are shifted left by 13.8x to define Omicron titres. The GMTs are indicated. Note that most of the Omicron titres are below the assay detection limit of 40. (See footnote 1 for a technical detail on the figure.)

### A.2 Neutralisation data

#### The Legacy Study

Vaccine effectiveness is modelled as a function of 50% neutralising antibody titres against SARS-CoV-2 variants. These data came from the Legacy Study study in the UK with Comirnaty-vaccinated volunteers from the UCLH and the Crick Institute. The study is described in Wall et al. [2021], and its anonymised data are available from its GitHub repository. The one-dose cohort included 186 participants; and the two-dose cohort, 160 (based on the study data on GitHub). Out of the 248 unique subjects, 98 participated in both cohorts. The participants’ sera were tested in live-virus neutralisation assays with the Wild Type; the D614G lineage; and the Alpha, Beta and Delta VOCs. The measurements from a participant were used to estimate her or his titre attaining 50% neutralisation of the variant. Example titres from the two-dose cohort are shown in Figure 3. An estimated titre above the assay detection limit of 2560 was recorded as 5120, and that below the limit of 40, as 10. Cases with no observed neutralisation were recorded as 5. (More details are in Wall et al. [2021] and Volkov et al. [2021].)

#### Titres against Omicron

To model neutralising titres against Omicron we could use the titres against the Wild Type [Khoury et al., 2021, Cromer et al., 2021]. However, as illustrated in Figure 3, the titres against Beta, an immediate predecessor, would approximate Delta titres better than Wild Type titres would. We therefore modelled Omicron titres as a fold of the Delta titres in the two-dose cohort. As an example, consider the Polymutant scenario with a 80x decrease versus the Wild Type. The median Delta titre in the two-dose cohort of Legacy was 5.8x less than the median Wild-Type titre [Wall et al., 2021]. Therefore Omicron titres after two doses were computed by multiplying the Delta titres by 13.8x, since 13.8 *×* 5.8 = 80; see^1^ Figure 4. Since no post-booster titres were available, we used the two-dose Delta titres to construct Omicron titres after boosting: the two-dose Omicron titres were multiplied by 3.3, the assumed fold increase from boosting.

### A.3 Effectiveness modelling

The model used for vaccine effectiveness is as follows. Let the vector ***t***_d,*ν*_ = (*t*_1_, …, *t*_*n*_) contain log10-transformed 50% neutralising titres against variant *ν* after d vaccine doses. There are *n* titres altogether, with titre *t*_*i*_ corresponding to participant *i*. For example, ***t***_d,*ν*_ could contain 157 titres sampled against Delta in the two-dose Legacy cohort, see Figure 3. This vector is the explanatory variable in the model for vaccine effectiveness,

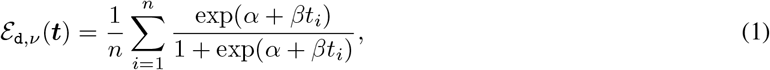

which is evaluated for dose count d and variant *ν*. The model combines data on a particular vaccine across doses and variants. Its parameters, *α* and *β*, are fixed for every d, *ν* and participant *i*. (This and similar models are widely used in vaccine literature, e.g. by Dunning [2006], Nauta et al. [2009] and Khoury et al. [2021].) Since vaccine effectiveness is a percent between 0 and 100, we assumed that the random errors about the model function followed a beta distribution, which is particularly suitable for response on a bounded interval. Consequently, the model was fitted with nonlinear beta regression, via maximum likelihood.

To predict symptomatic effectiveness against Omicron, we fitted the model to 32 Comirnaty effectiveness estimates from 14 published studies across 10 countries; see Volkov et al. [2021] for details. The estimates were obtained against specific variants in once or twice-vaccinated populations, aged over 18. The estimates for two Comirnaty doses corresponded to three-five months starting two-three weeks after the second dose. We therefore assumed the same timing for predicted effectiveness against Omicron. The explanatory variable was evaluated at the titre samples against D614G, Alpha, Beta and Delta from the Legacy study with one and two-dose Comirnaty cohorts. The resulting model fit is plotted in Figure 5 against the geometric mean titre (GMT) per per dose/variant combination. The predictions in this paper assumed equal variances of effectiveness estimates. All the effectiveness predictions (not shown) for Omicron under unequal variances were slightly larger than the corresponding predictions reported here. Unequal variances were investigated in Volkov et al. [2021] and are implemented in the software package.

**Figure 5:**
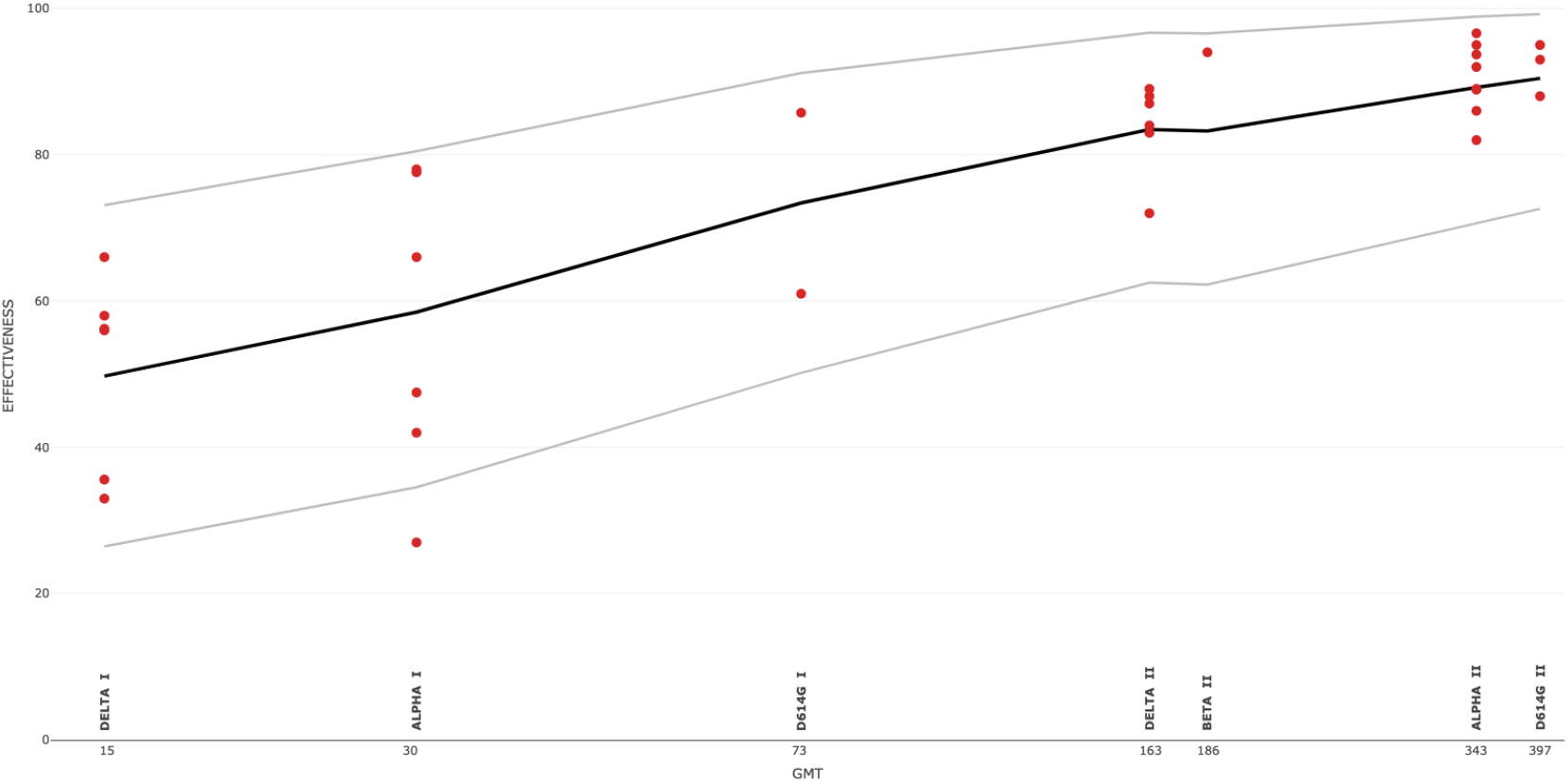
Model function (1) fitted to 32 effectiveness estimates and Legacy neutralisation data on the indicated variants and doses. The 95% prediction bands are shown. The x-axis is on a log10 scale.

### A.4 Model assessment

This section presents a key issue with the modelling behind the predictions, and argues why they can be informative regardless. There is growing evidence that the assumed explanatory variable — 50% neutralising antibody titres — is strongly correlated with Covid vaccine efficacy and effectiveness (Feng et al. [2021], Gilbert et al. [2021], Khoury et al. [2021]). A crucial point for our modelling, however, is that the Legacy population, which provided the titres, differs from the populations that provided the effectiveness estimates, and hence the “neutralisation population” titres may differ from the “effectiveness population” titres. Such data were simply not available from the same population. In fact, they may be impractical to collect, for instance, when neutralising titres are determined in live-SARS-CoV-2 assays, and could be difficult to compare across studies. (There are other issues, considered in Volkov et al. [2021], such as disparate definitions of vaccine effectiveness between studies.) Therefore we followed the same data-driven approach, as in Khoury et al. [2021], Cromer et al. [2021] and Earle et al. [2021], by using effectiveness and neutralisation data from different populations.

The resulting predictions, including the ones in this paper, are subject to potential biases due to population differences. Still, we cautiously conclude that the Legacy population can serve as a practical model for effectiveness populations and enable useful predictions of vaccine effectiveness. First, some of the biases can be mitigated by matching effectiveness populations with the Legacy population, which is relatively large and diverse, for instance, in terms of age, ranging between 20 and 70. (Beyond the assumption of adult populations, no matching was done for the Omicron predictions.) Second, the Legacy use of a single, high-quality laboratory and of live-virus assays avoids errors due to inter-laboratory variation and pseudovirus assays. Third, what is likely to matter for modelling is not the actual titres, but their change between variants, that is, the “folds”. Although the former can differ widely across individuals, the latter may differ less, because of common biological processes and correlations within an individual. Furthermore, most, if not all, neutralisation studies have relatively small, non-random populations, with the Legacy’s possibly the largest; and yet they produce valuable results in terms of neutralisation folds. The intuition is that these folds are informative of vaccine effectiveness despite the non-representative populations studied. Our modelling captures the fold differences in the Legacy population and quantifies them as vaccine effectiveness, and so enhances the usual intuitive assessment of neutralisation folds. Fourth, elements were added, such as the beta distribution for effectiveness, that make predictions more accurate compared to other approaches. Unlike the framework here, even the purpose-designed studies, such as Feng et al. [2021] and Gilbert et al. [2021], of correlates of vaccine protection did not examine data after the first dose. But such data could improve the models, as could be seen in Figure 5, especially for VOCs like Omicron, which push towards the first-dose regions. These and other arguments (e.g. that the function (1) mitigates some of the population differences) are elaborated in Volkov et al. [2021].

Another argument for the proposed framework is that it produced remarkably accurate predictions for pre-Omicron variants. This was even without population matching and other statistical adjustments, which could increase the accuracy further. For instance, Figure 6, redrawn from Volkov et al. [2021], shows “leave-one-variant-out” predictions obtained by fitting the model without data on the variant, and then evaluating the fitted model at the neutralising titres against the variant. The largest prediction error overall was 3.2, in percent, for one Comirnaty dose against Alpha. The errors for two doses were -1.6, -0.5 and -0.6 against D614G, Alpha and Delta, respectively, with the “minus” denoting underperdiction. The 95% prediction bands covered every estimate with a single exception of one-dose effectiveness against Alpha. Other predictions in considered in that paper were also accurate, including those

**Figure 6:**
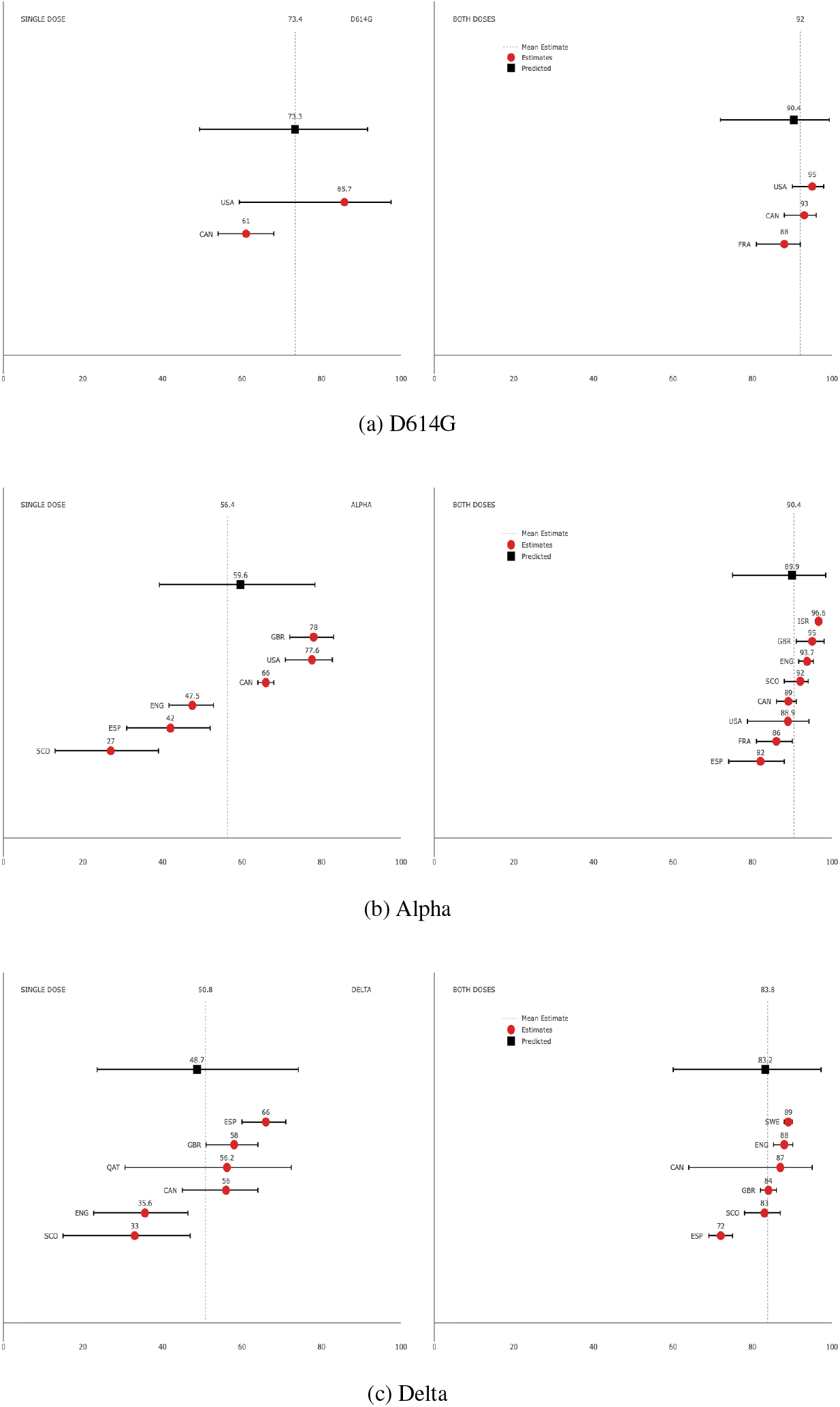
Predicted symptomatic effectiveness of Comirnaty against SARS-CoV-2 variants after one or two primary doses. The origin countries for Comirnaty effectiveness studies are indicated. See Volkov et al. [2021] for the effectiveness data and other details.

- under unequal variances of effectiveness estimates;
- using 12 effectiveness estimates from the UK studies only, versus 32 in the full data set. Such “leave-one-variant-out” prediction for Alpha only used six effectiveness estimates for Delta; similarly, for Delta;
- in subsets of study participants (e.g. 98 participants, out of 248, who were in both cohorts);
- of Vaxzevria vaccine effectiveness either directly from Vaxzevria data or indirectly from the model fitted to Comirnaty data.

See Volkov et al. [2021] for more details and comparisons with effectiveness predictions made in Cromer et al. [2021] and Chen et al. [2021].

Although Omicron predictions cannot be assessed yet, its effectiveness could closely follow the trend in Figure 5, which shows a clear, almost linear relationship (on a log-scale) for the doses and variants considered.

## Data Availability

All data produced are available online from a public GitHub repository.

https://github.com/XitificOpenSource/Pfizer_BioNTech_BNT162b2_vaccine_effectiveness_vs_Omicron

## Data and code availability

All data and R code used for this paper are freely available from the paper’s GitHub repository

## Acknowledgements

I am grateful to Rosemary Bailey, Ian Bartlett, Jason Fogaros, Alexander Gray and Andrew Langley for their support during this research, and to Andrey Kajava for fruitful discussions.

## Footnotes

1 To aid visualisation, the censored titres were shifted towards their respective detection limits. The assumed difference between the Wild Type and Delta titres was defined as 5.8x (5.0, 6.9) reported in Wall et al. [2021]. This is a bootstrap estimate of the ratio of the median Wild Type titre to the median Delta titre, and is slightly larger than 892.8*/*162.6 *≈* 5.5, the ratio of the respective geometric mean titres in the Figure.

## References

ACI. Living evidence - SARS-COV-2 variants, Nov 2021. URL https://aci.health.nsw.gov.au/covid-19/critical-intelligence-unit/sars-cov-2-variants.

Robert L. Atmar, Kirsten E. Lyke, Meagan E. Deming, Lisa A. Jackson, et al. Heterologous SARS-CoV-2 booster vaccinations – preliminary report. October 2021. doi:10.1101/2021.10.10.21264827. URL https://doi.org/10.1101/2021.10.10.21264827.

Anna Bernasconi, Pietro Pinoli, Ruba Al Khalaf, Tommaso Alfonsi, et al. Report on Omicron Spike mutations on epitopes and immunological/epidemiological/kinetics effects from literature. http://Virological.org, 2021. URL https://virological.org/t/report-on-omicron-spike-mutations-on-epitopes-and-immunological-epidemiological-kinetics-effects-from-literature/770.

Sandile Cele, Laurelle Jackson, Khadija Khan, David Khoury, et al. SARS-CoV-2 Omicron has extensive but incomplete escape of Pfizer BNT162b2 elicited neutralization and requires ACE2 for infection. December 2021. URL https://www.ahri.org/wp-content/uploads/2021/12/MEDRXIV-2021-267417v1-Sigal.pdf.

Xinhua Chen, Andrew S. Azman, Wanying Lu, Ruijia Sun, et al. Prediction of vaccine efficacy of the delta variant. August 2021. doi:10.1101/2021.08.26.21262699. URL https://doi.org/10.1101/2021.08.26.21262699.

Lynsey Chutel, Richard Pérez-Peña, and Emily Anthes. Omicron is fast moving, but perhaps less severe, early reports suggest. The New York Times, 2021. URL https://www.nytimes.com/2021/12/06/world/africa/omicron-coronavirus-research-spread.html.

Deborah Cromer, Megan Steain, Arnold Reynaldi, Timothy E Schlub, et al. Neutralising antibody titres as predictors of protection against SARS-CoV-2 variants and the impact of boosting: a meta-analysis. The Lancet Microbe, November 2021. doi:10.1016/s2666-5247(21)00267-6. URL https://doi.org/10.1016/s2666-5247(21)00267-6.

Andrew J. Dunning. A model for immunological correlates of protection. Statistics in Medicine, 25(9):1485–1497, 2006. doi:10.1002/sim.2282. URL https://doi.org/10.1002/sim.2282.

Kristen A. Earle, Donna M. Ambrosino, Andrew Fiore-Gartland, David Goldblatt, et al. Evidence for antibody as a protective correlate for COVID-19 vaccines. Vaccine, May 2021. doi:10.1016/j.vaccine.2021.05.063. URL https://doi.org/10.1016/j.vaccine.2021.05.063.

FDA. Emergency use authorization (eua) for an unapproved product review memorandum, 2021. URL https://www.fda.gov/media/152432/download.

Shuo Feng, Daniel J. Phillips, Thomas White, Homesh Sayal, et al. Correlates of protection against symptomatic and asymptomatic SARS-CoV-2 infection. Nature Medicine, September 2021. doi:10.1038/s41591-021-01540-1. URL https://doi.org/10.1038/s41591-021-01540-1.

Peter B. Gilbert, David C. Montefiori, Adrian McDermott, Youyi Fong, et al. Immune correlates analysis of the mRNA-1273 COVID-19 vaccine efficacy trial. August 2021. doi:10.1101/2021.08.09.21261290. URL //doi.org/10.1101/2021.08.09.21261290.

David S. Khoury, Deborah Cromer, Arnold Reynaldi, Timothy E. Schlub, et al. Neutralizing antibody levels are highly predictive of immune protection from symptomatic SARS-CoV-2 infection. Nature Medicine, May 2021. doi:10.1038/s41591-021-01377-8. URL https://doi.org/10.1038/s41591-021-01377-8.

Sharon LaFraniere and Noah Weiland. Blood samples of people who received two pfizer doses showed a 25-fold reduction in antibody levels against omicron. The New York Times, 2021. URL https://www.nytimes.com/live/2021/12/08/world/omicron-variant-covid#pfizer-says-blood-samples-showed-a-third-dose-of-its-vaccine-provides-significant-protection-against-omicron.

Yuying Lu, Jin Wang, Qianlin Li, Huan Hu, Jiahai Lu, and Zeliang Chen. Advances in neutralization assays for SARS-CoV-2. Scandinavian Journal of Immunology, 94(3), June 2021. doi:10.1111/sji.13088. URL https://doi.org/10.1111/sji.13088.

Jozef J.P. Nauta, Walter E.P. Beyer, and Albert D.M.E. Osterhaus. On the relationship between mean antibody level, seroprotection and clinical protection from influenza. Biologicals, 37(4):216–221, August 2009. doi:10.1016/j.biologicals.2009.02.002. URL https://doi.org/10.1016/j.biologicals.2009.02.002.

Pfizer/BioNTech. Pfizer and biontech provide update on omicron variant, 2021. URL https://www.businesswire.com/news/home/20211208005542/en/.

Fabian Schmidt, Yiska Weisblum, Magdalena Rutkowska, Daniel Poston, et al. High genetic barrier to SARS-CoV-2 polyclonal neutralizing antibody escape. Nature, September 2021. doi:10.1038/s41586-021-04005-0. URL https://doi.org/10.1038/s41586-021-04005-0.

Oleg Volkov, Svetlana Borozdenkova, and Alexander Gray. Predicting the effectiveness of covid-19 vaccines from SARS-CoV-2 variants neutralisation data. September 2021. doi:10.1101/2021.09.06.21263160. URL https://doi.org/10.1101/2021.09.06.21263160.

Emma C Wall, Mary Wu, Ruth Harvey, Gavin Kelly, et al. Neutralising antibody activity against SARS-CoV-2 VOCs B.1.617.2 and B.1.351 by BNT162b2 vaccination. The Lancet, 397(10292):2331–2333, June 2021. doi:10.1016/s0140-6736(21)01290-3. URL https://doi.org/10.1016/s0140-6736(21)01290-3.

Pengfei Wang, Manoj S. Nair, Lihong Liu, Sho Iketani, et al. Antibody resistance of SARS-CoV-2 variants b.1.351 and b.1.1.7. Nature, 593(7857):130–135, March 2021. doi:10.1038/s41586-021-03398-2. URL https://doi.org/10.1038/s41586-021-03398-2.

C. Zimmer. New virus variant stokes concern but vaccines still likely to work. The New York Times, 2021. URL https://www.nytimes.com/2021/11/26/health/omicron-variant-vaccines.html?smid=url-share.

